# Effectiveness of mass screening for severe acute malnutrition using mid-upper arm circumference: a prospective regression discontinuity design

**DOI:** 10.1101/2025.04.18.25326071

**Authors:** Huiyu Hu, Mamadou Ouattara, Mamadou Bountogo, Valentin Boudo, Thierry Ouedraogo, Clarisse Dah, Elodie Lebas, Till Bärnighausen, Benjamin F. Arnold, Kieran S. O’Brien, Thomas M. Lietman, Ali Sié, Catherine E. Oldenburg

## Abstract

**Background:** Severe Acute Malnutrition (SAM) is a major cause of morbidity and mortality in children under five, particularly in sub-Saharan Africa. The World Health Organization (WHO) recommends using mid-upper arm circumference (MUAC) for community-based screening due to its simplicity and cost-effectiveness. This study assesses the impact of MUAC-based screening and referral by community health workers (CHWs) on nutritional status and all-cause mortality in Burkina Faso.

**Methods:** A prospective regression discontinuity (RD) analysis was nested within the Community Health with Azithromycin Treatment (CHAT) trial, a cluster-randomized controlled trial in Nouna District, Burkina Faso. Children aged 6-59 months were screened using MUAC, and those with MUAC < 11.5 cm were referred for SAM treatment. The effect of referral on subsequent MUAC measurements and 6-month all-cause mortality was analyzed using data near the 11.5 cm threshold. Bandwidths were selected using the Imbens-Kalyanaraman (IK) algorithm to optimize bias-variance tradeoff. Generalized linear mixed-effects models were applied for analysis.

**Results:** Out of 65,554 children screened, 226 (0.34%) were referred for SAM. Using the IK-selected bandwidth for MUAC, no significant effects on 6-month MUAC or mortality were observed. The mean MUAC difference was 0.21 cm (95% CI: −0.22 – 0.65 cm), and the mortality odds ratio was 0.95 (95% CI: 0.03 – 31.37). Sensitivity analyses using broader bandwidths yielded consistent results.

**Conclusions:** MUAC-based screening and referral did not significantly improve nutritional status or reduce mortality. Improving SAM referral mechanisms and integrating community health support are necessary to improve health outcomes for children with SAM.

**Key Message:**

- We evaluated whether MUAC-based community screening and referral for Severe Acute Malnutrition (SAM) improved nutritional status and reduced mortality among young children in rural Burkina Faso using a prospective regression discontinuity design.
- MUAC screening and referral alone did not significantly affect subsequent nutritional outcomes or all-cause mortality.
- These findings highlight the importance of strengthening referral completion and integrated community-based care to ensure that screening translates into meaningful improvements in child health.

## Introduction

Globally, severe acute malnutrition (SAM) is a leading contributor to morbidity and mortality in children under five years of age, (1) especially in sub-Saharan Africa and South Asia. In Burkina Faso, malnutrition presents a significant challenge, with data indicating that 18% of children under the age of five are underweight (low weight for age), 9% are wasted (low weight relative to height), and 25% are stunted (low height for age). (2) In the 2013 guideline for the management of SAM, the World Health Organization (WHO) advised the use of mid-upper arm circumference (MUAC) for community-based screening by community health workers (CHWs) for severe and moderate acute malnutrition in children under five years old, using cut-offs of <11.5cm for SAM and <12.5cm for moderate acute malnutrition (MAM). (3) MUAC is a simple, quick, and cost-effective tool that can be used at the community level to screen children for acute malnutrition and facilitate referral to nutritional programs. Its simplicity enables it to be used by CHWs and laypersons after minimal training, (4) facilitating widespread screening in resource-limited settings. In many nutritional programs, MUAC has been implemented as the sole criterion for admission and discharge, leading to good rates of recovery and weight gain. (5,6) A previous study provided evidence indicating that CHWs have a high capacity to accurately diagnose SAM using MUAC measurements. (7) While MUAC has been widely adopted to assess SAM in community-based management of acute malnutrition (CMAM) programs, the effectiveness of mass screening and referral for treatment for SAM by CHWs is unclear.

Regression discontinuity (RD), a quasi-experimental design, allows for causal evaluation of interventions that use threshold rules that are used to assign interventions based on the measurement of a continuous variable (the “running variable”). (8–10) In the case of community screening for acute malnutrition, MUAC is a continuous variable measured with noise that is used to determine whether a child is referred for treatment of SAM based on the threshold of MUAC < 11.5 cm. In this study, we evaluated the effectiveness of CHW screening and referral to nutritional programs for SAM on nutritional status and all-cause mortality, utilizing a prospective RD design nested within a large cluster-randomized controlled trial.

## Methods

### Parent trial design

The Community Health with Azithromycin Treatment (CHAT) trial was a cluster randomized trial evaluating twice yearly mass distribution of a single, oral dose of azithromycin compared to placebo for prevention of all-cause mortality in children aged 1-59 months. Complete methods and the primary outcome for the trial have been previously reported. (11,12) In brief, 341 communities in Nouna District, Burkina Faso were 1:1 randomized to biannual mass azithromycin distribution or placebo to children aged 1-59 months. A door-to-door enumerative census was conducted every 6 months, during which the vital status of each child was assessed and children received study treatment (azithromycin or placebo). The study was conducted from August 2019 until February 2023, corresponding to 6 rounds of census and treatment.

The study was reviewed and approved by the Institutional Review Board at the University of California, San Francisco and the Comité d’Ethique pour la Recherche en Santé in Ouagadougou, Burkina Faso. Written informed consent was obtained from at least one guardian of each child enrolled in the study.

### Study setting

CHAT was conducted in Nouna District, Burkina Faso, a rural and mostly agrarian district in northwestern Burkina Faso near the Mali border. Burkina Faso is located in the Sahel, which experiences highly seasonal rainfall from July through October, followed by an annual harvest in November-December. Food insecurity, and thus acute malnutrition, are typically highest during the rainy season while crops are growing. The rainy season also corresponds with the higher malaria transmission season.

### Communities and participants

All communities in Nouna District were eligible for inclusion in the trial. After randomization, several communities were lost due to escalating political insecurity occurring in the study area. (12) All children in the eligible age range (1 to 59 months) were eligible for treatment during each twice-yearly treatment round. Children were censused up to 65 months of age to evaluate mortality in the 6-month period following each round of treatment administration.

Because MUAC-based cutoffs are only validated and used for children ≥6 months of age, this analysis was restricted to children who were 6-59 months of age at the time of first MUAC measurement.

### Running variable measurement

During each census, MUAC-based screening for SAM was conducted by trained study census workers using standard MUAC tapes. Census workers used a string to identify the midpoint of the child’s upper arm, and then measured the circumference using the MUAC tape. Numeric MUAC measurements were entered by the enumerator into a custom-designed mobile data collection application designed specifically for the trial. The application was flagged any child with a MUAC measurement below 11.5 cm, instructing the enumerator to refer the child to a primary healthcare facility for diagnosis and enrollment in the nutritional program for treatment of SAM. However, due to limitations linking census data with health facility records, the trial did not track whether caregivers sought recommended care.

### Outcomes

We assessed the effect of screening and referral to the nutritional program on two outcomes: 1) subsequent MUAC measurement and 2) all-cause mortality. Both MUAC and all-cause mortality were measured during each twice-yearly census round. Children were followed longitudinally up to 65 months of age, although children over 59 months of age did not receive an additional round of treatment. Therefore, a 59-month-old child who received MUAC measurements would be included in the study until they were 65 months of age. MUAC measurements were obtained as previously described. Deaths were included in the study if a child was recorded as alive and living in the household during one census and died prior to the next census.

## Statistical analysis methods

### Regression Discontinuity Design

To evaluate the impact of MUAC screening on follow-up MUAC measurements and mortality, a RD design was employed, using MUAC as the running variable with a threshold of 11.5 cm for intervention eligibility (referral to the nutritional program). The RD design is a quasi-experimental approach used to identify the causal effects of interventions by taking advantage of arbitrary threshold rules. Due to a considerable amount of noise, children with a MUAC measurement of 11.4 cm and 11.6 cm are likely very similar in their risk of outcomes. Thus, in a narrow range around the threshold, exchangeability can be a reasonable assumption, and causal effects can be identified by comparing participants with measurements immediately above and below the threshold. In this study, the RD design allows for the estimation of the causal effect of MUAC screening on children who are near the cutoff for SAM referral (11.5 cm), offering an opportunity to compare outcomes for children just above and just below the eligibility threshold.

Bandwidth selection in RD involves a bias-variance tradeoff. A wider bandwidth decreases the variance of the estimates by increasing the number of observations but also increases the bias because it relies on correctly specifying the functional form of the running variable above and below the threshold. Conversely, narrower bandwidths minimize bias but reduce statistical power due to smaller sample sizes. To find an optimal balance between bias and variance, the Imbens-Kalyanaraman (IK) algorithm was implemented to select the optimal bandwidths for MUAC for each outcome, enhancing the precision and validity of the estimates. (13) Additionally, three bandwidths (10.5–12.5 cm, 9.5–13.5 cm, and 8.5–14.5 cm) were used for robustness checks and sensitivity analyses, ensuring the consistency and reliability of the study’s findings.

### Regression Discontinuity Assumptions

In our study, assumptions for causal inference in RD were assessed as follows: (9,10)

1. The MUAC served as our assignment variable, measured and reported continuously.
2. The intervention eligibility (referred or not referred to the nutritional program) was determined by a clearly defined cut-off value of 11.5 cm in MUAC measurements.
3. The assignment variable, MUAC, measured continuously near the cut-off value, was assumed to be not influenced by external factors.
4. MUAC and vital status were recorded for all eligible children during the census, regardless of their exposure to the referral intervention.
5. We verified the presence of individuals both above and below the MUAC threshold within our population, ensuring a viable comparison across the intervention cutoff.
6. To ensure the precision of our analysis, we undertook robustness checks that involved employing models for the assignment variable, particularly MUAC, across different bandwidths.

For the analysis, generalized linear mixed-effects models were utilized, incorporating three different bandwidths selected from IK algorithm. Specifically, the regression model was fitted in following forms:

1) 6-month followup MUAC as outcome:

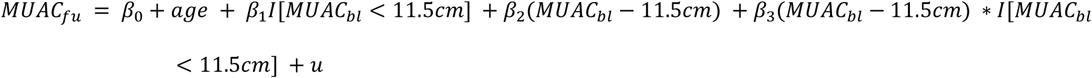

2) Vital status at 6-month as outcomes

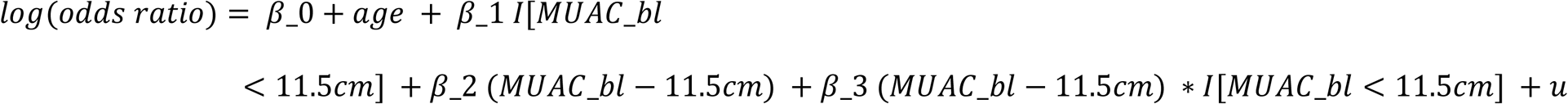

Where 𝐼[𝑀𝑈𝐴𝐶_𝑏𝑙_ < 11.5𝑐𝑚] indicates whether a child was referred to the nutritional program or not, 𝛽_1_is the difference in the cutoff, which is equivalent to the effect of eligibility of nutritional program referral as defined by the threshold, 𝛽_2_ is the slope of the line below the threshold, 𝛽_2_ + 𝛽_3_ is the slope of the line above the threshold and 𝑢 represents the random effects term, accounting for unobserved heterogeneity at the individual level and the fact that one child could contribute to multiple treatment phases. Considering that a child’s age is associated with mortality and MUAC, the baseline age was included in the models to reduce the variance. Analyses were completed in R version 4.2.

As a sensitivity analysis, we additionally controlled for the study treatment arm (azithromycin vs. placebo) as well as other baseline covariates, including distance to the nearest CSPS (health facility), and the number of children under 5 years old in the household. A squared term for the running variable was also included in the regression model to evaluate the robustness of the results.

## Result

A total of 68,246 children 1-59 months were enrolled in the CHAT study. After excluding children younger than 6 months for WHO-recommended age range for MUAC screening and those with a MUAC below 6 cm as outliers, 226 out of 65,554 (0.34%) children were diagnosed with SAM and referred at least once during the CHAT study period (Supplementary Figure 1).

Using the IK algorithm, we identified optimal bandwidths for each outcome. For MUAC at follow-up, the optimal bandwidth was 0.70 cm (10.80–12.20 cm), with 158 observations below the cutoff (met the criteria for SAM referral) and 2,743 above. For follow-up mortality, the optimal bandwidth was 0.62 cm (10.88–12.12 cm), with 148 observations below the cutoff and 2,480 above. Participants’ characteristics are shown in Table 1. Supplementary Figure 2 displays the MUAC distribution, demonstrating continuity at the MUAC cutoff.

**Table 1.** Baseline Characteristics.

|  | 10.80 - 12.20 cm <sup>1</sup> | 10.88 - 12.12 cm <sup>2</sup> |
| --- | --- | --- |
| **Number of records** | 2,901 | 2,628 |
| **Number of children** | 2,654 | 2,409 |
| **Baseline Age (in month, mean [SD])** | 18.7 (12.1) | 18.7 (12.2) |
| **Baseline MUAC (in month, mean [SD])** | 11.9 (0.3) | 11.9 (0.2) |
| **Number of SAM referrals (N [%])** | 158 (5.4%) | 148 (5.6%) |
| **Number of SAM children (N [%])** | 156 (5.9%) | 146 (6.1%) |
| **Number of female (N [%])** | 1489 (56.1%) | 1333 (55.3%) |

<sup>1</sup> The optimal bandwidth for Mean MUAC at 6 months, selected using the IK algorithm.
<sup>2</sup> The optimal bandwidth for Mortality at 6 months Odds Ratio selected using the IK algorithm.

From the RD analysis to estimate discontinuity at the MUAC cut-off of 11.5 cm, we were unable to demonstrate evidence of an effect of screening and referral for SAM on all-cause mortality or nutritional status as measured by MUAC at 6 months, with results broadly consistent across bandwidths in sensitivity analysis (Supplementary Table 1). Table 2 presents the estimation of the causal effect of MUAC-based screening on MUAC and mortality using the IK-selected bandwidths, as well as pre-specified bandwidths for robustness checks. Within the IK-selected bandwidth of 10.80–12.20 cm for MUAC follow-up, the mean difference in MUAC at 6 months was 0.21 cm (95% CI: −0.22 to 0.65 cm). Similarly, for mortality at 6-month follow-up, within the IK-selected bandwidth of 10.88–12.12 cm, the odds ratio (OR) was 0.95 (95% CI: 0.03 to 31.37), indicating no significant difference between children referred to the nutritional program (below the threshold) and those not referred (above the threshold). Supplementary Table 2 and 3 shows the estimation of causal effects of SAM screening, incorporating adjustments for covariates (baseline age, distance to CSPS, number of children (under 5) in household and treatment arm) and including a squared term in the regression model for robustness checks. Both sensitivity analyses yielded consistent results, indicating that there was no significant difference in mortality or MUAC outcomes referred to the nutritional program and those not referred.

**Table 2.** Model outcomes Using IK-Selected Bandwidth.

| Outcome | Estimate (95% CI) | P-value |
| --- | --- | --- |
| Mean MUAC at 6 months (cm) <sup>1</sup> | 0.21 (-0.22 – 0.65) | 0.339 |
| Mortality Odds Ratio (OR) <sup>2</sup> | 0.95 (0.03 – 31.37) | 0.978 |

<sup>1</sup> The optimal bandwidth for Mean MUAC at 6 months was 0.70 cm (10.80–12.20 cm), selected using the IK algorithm.
<sup>2</sup> The optimal bandwidth for Mortality at 6 months Odds Ratio was 0.62 cm (10.88–12.12 cm), selected using the IK algorithm.

## Discussion

In this prospective RD analysis nested in a cluster randomized trial, we did not find evidence that MUAC-based screening and referral to nutritional programs improved MUAC measures 6 months after screening. However, confidence intervals in particular for the mortality endpoint were wide, as SAM was less common than expected during the study period and mortality is a rare event. At the same time, since MUAC is continuous, this outcome was better powered than mortality, and estimates consistently showed no evidence of effect. While children with SAM have a higher mortality rate than children without SAM and this endpoint is an important indicator for SAM treatment programs, (3,14) programs monitor response to treatment with MUAC, and children who are admitted to nutritional programs are considered to have recovered when their MUAC increases to > 12.5 cm.

3) Lack of an effect of screening and referral to the nutritional program on subsequent MUAC measurements and mortality for children with SAM may suggest inadequate referral linkage, indicating families affected by SAM require additional support. CHAT lacked mechanisms such as follow-up visits to ensure SAM referral completion, and no data were collected on whether referred children accessed care. Future studies should evaluate both screening effectiveness and referral completion to improve SAM outcomes.

The findings of this analysis underscore the challenges in connecting referred SAM children to appropriate care and raises questions about the effectiveness of screening alone in improving SAM outcomes. While MUAC screening is widely endorsed and used globally, its success depends on the presence of robust downstream referral and treatment pathways. A previous study estimated that the average coverage of CMAM programs was only 38.3% in sub-Saharan Africa, identifying several obstacles such as a lack of awareness about malnutrition and the programs themselves, along with geographical and economic access barriers to health facilities. (15) In addition, structural barriers, including poverty and lack of access to healthcare can limit caregivers’ ability to follow through with referrals and provide adequate care for their children. For example, an evaluation of a CMAM program in Uganda, caregivers faced significant economic and social constraints that hindered their ability to provide adequate care. (16) These structural barriers must be addressed to make interventions more effective in contexts of extreme poverty in area like rural Burkina Faso.

To overcome these challenges, previous studies have identified effective intervention strategies. For example, the integrated community case management (iCCM) approach involves integrating SAM treatment with the existing curative tasks of CHWs, thereby enhancing treatment accessibility and potentially improving recovery outcomes, particularly for children in rural areas. (17) Training mothers/caregivers to regularly screen for SAM using MUAC presents a cost-effective strategy that enhances coverage, helps early detection of SAM, and promotes engagement in seeking SAM treatment. (18,19)

Additionally, the optimal indicator for detecting acute malnutrition may still require further investigation. MUAC is widely used for SAM screening due to its simplicity and cost effectiveness. However, ongoing debates in the literature suggest that relying solely on the current WHO-recommended MUAC cutoff without incorporating weight-for-height (WHZ) as a screening tool may lead to an underestimation of severe acute malnutrition cases. (20,21) Yet, WHZ measurement is more complex, requiring calibrated scales and trained personnel, making it impractical for routine use in community-based settings. Future research should explore integrating MUAC with other indicators to improve the identification of malnutrition while considering the practical constraints of community-based programs.

## Limitations

The study faced several limitations that could impact the interpretation and generalization of findings. First, fewer SAM cases and lower-than-expected mortality led to wide confidence intervals for the mortality endpoint. RD analyses are often restricted to relatively narrow bandwidths, which further limits statistical power. As a result, the mortality endpoint was very underpowered in this analysis, and results were consistent with a wide range of effects. The study did not collect data on whether referred children were linked to nutritional programs, leaving gaps in understanding whether those identified through MUAC screening ultimately received care or what their outcomes were. Results of this study should not be interpreted as evaluating the effectiveness of the nutritional programs themselves, but rather of screening and referral to nutritional programs. Another significant limitation was the absence of a mechanism within the study to screen and refer children with moderate acute malnutrition (MAM; MUAC < 12.5 cm), which means our study could not assess the effectiveness of the screening and referral process for children who fall into this category, which represents a crucial aspect of comprehensive malnutrition management. Although children with MAM have increased risk of progression to SAM and mortality, infrastructure for provision of MAM treatment does not exist as it does for SAM. The CHAT trial was a large, simple trial, and as a result we were unable to collect data on some outcomes, such as WHZ, that may have provided additional information on the effect of screening and referral for SAM. Finally, the results of this analysis may not be generalizable to settings with different distributions of acute malnutrition and child mortality. Further evaluations of the effectiveness of mass screening and referral to nutritional programs are needed, particularly in settings with a higher prevalence of SAM. These results highlight the need for adequately powered evaluations to understand the impact of MUAC-based screening and referral on child health outcomes, including nutrition and mortality, and other relevant indicators.

## Conclusion

We found no evidence of an effect of mass screening using MUAC and referral to nutritional programs for SAM on subsequent nutritional status or all-cause mortality in this prospective RD design analysis. However, the prevalence of SAM was low, leading to uncertainty in estimates, especially for analyses of mortality. The results of this study suggest that current screening and referral mechanisms for SAM are not effectively improving outcomes for children with SAM, and additional health systems strengthening that includes, for example, enhanced linkage to nutritional programs via CHWs or other peer navigators, is needed.

**Figure 1.**
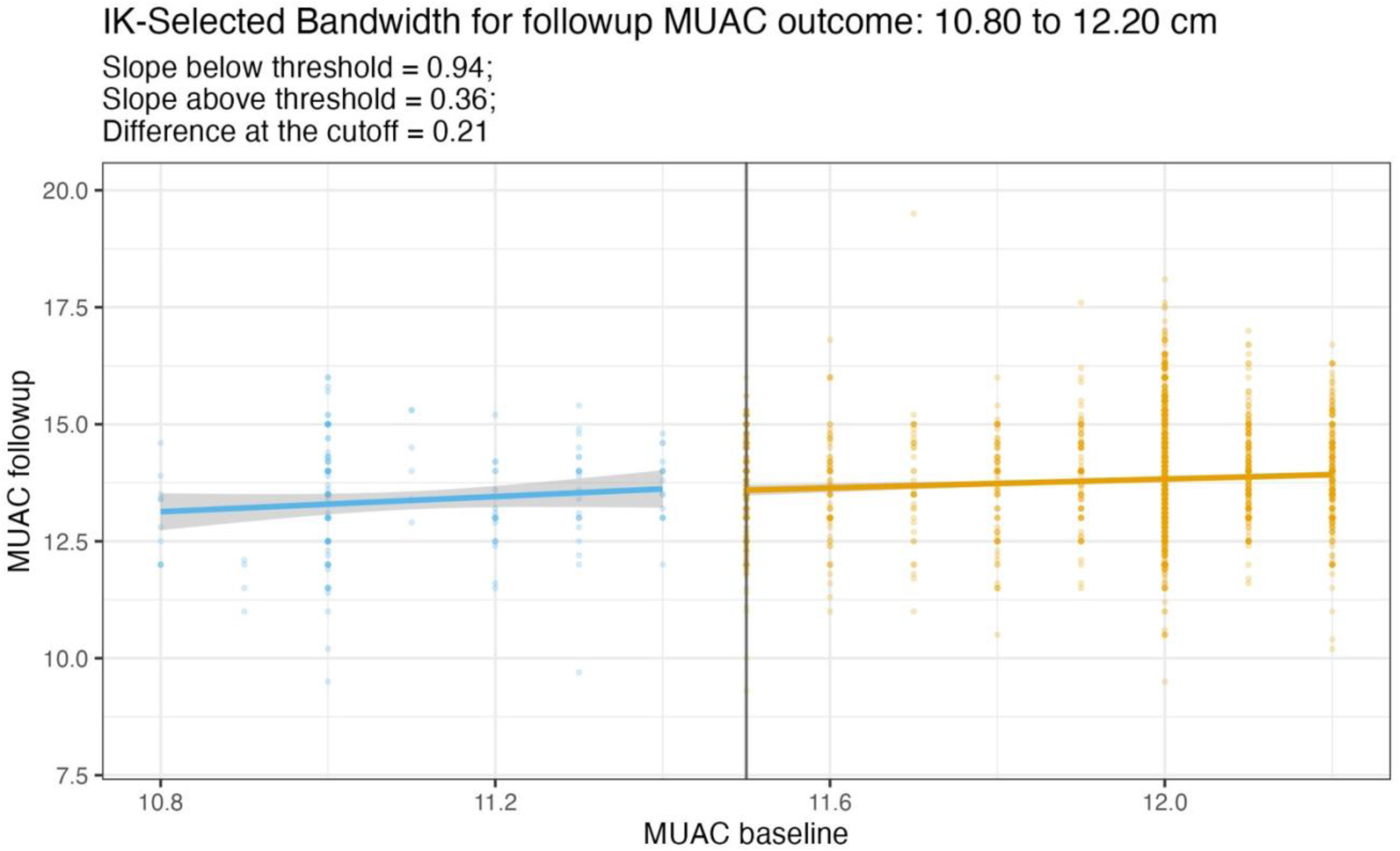
Regression discontinuity results for MUAC at 6 months.

## Data Availability

All data produced are available online at OSF

https://osf.io/4eg83/

## Acknowledgements

HH: study design, study implementation, data analysis, critical review, drafting manuscript. MO: study design, study implementation, supervision, critical review. MB: study design, study implementation, supervision, critical review. VB: study design, study implementation, data management, critical review. TO: study design, study implementation, supervision, critical review. CD: study implementation, supervision, critical review. EL: study design, study implementation, supervision, critical review. TB: study design, critical review, supervision, critical review. BFA: study design, study implementation, data analysis, critical review. KSO: study design, study implementation, critical review. TML: study design, study implementation, supervision, critical review. CEO: study design, study implementation, supervision, drafting manuscript.

## Data Availability

Data underlying all analyses are publicly available via the trials’ data repositories.

## Abbreviations

SAM: Severe Acute Malnutrition
MUAC: Mid-Upper Arm Circumference
WHZ: Weight-for-Height Z-score
WAZ: Weight-for-Age Z-score
CMAM: Community-based Management of Acute Malnutrition
CHWs: Community Health Workers
RD: Regression Discontinuity
WHO: World Health Organization
MAM: Moderate Acute Malnutrition

