## Supplementary Figure 1-2, Supplementary Table 1-3 for "Effectiveness of mass screening for severe acute malnutrition using mid-upper arm circumference: a prospective regression discontinuity design"

### Supplementary Figure 1. Participant Flow Chart

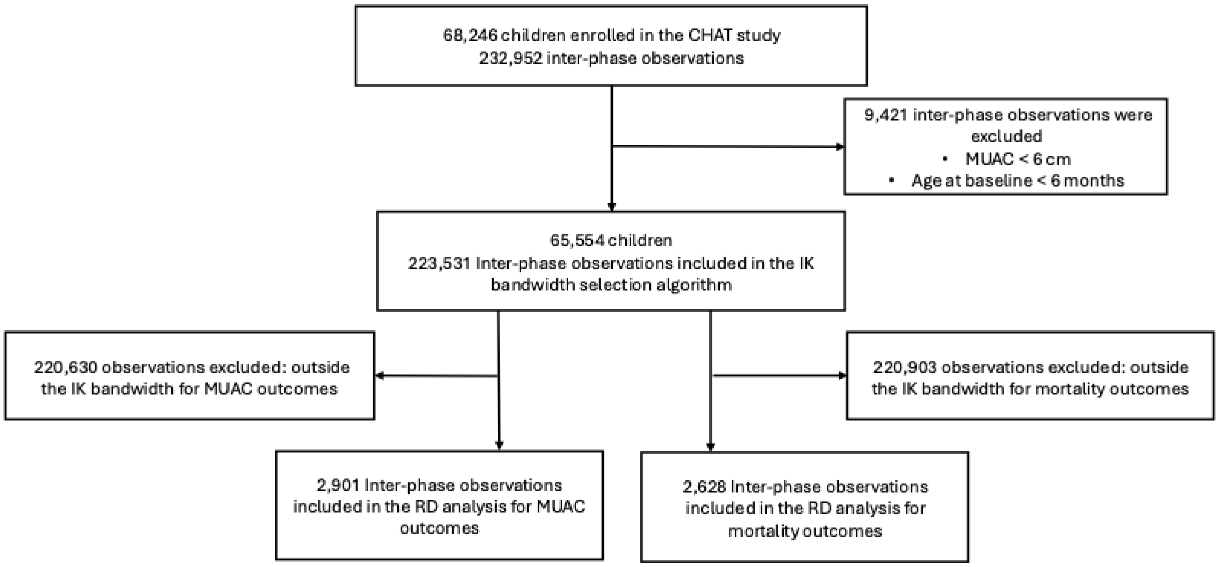

**Supplementary Figure 2. MUAC distribution**

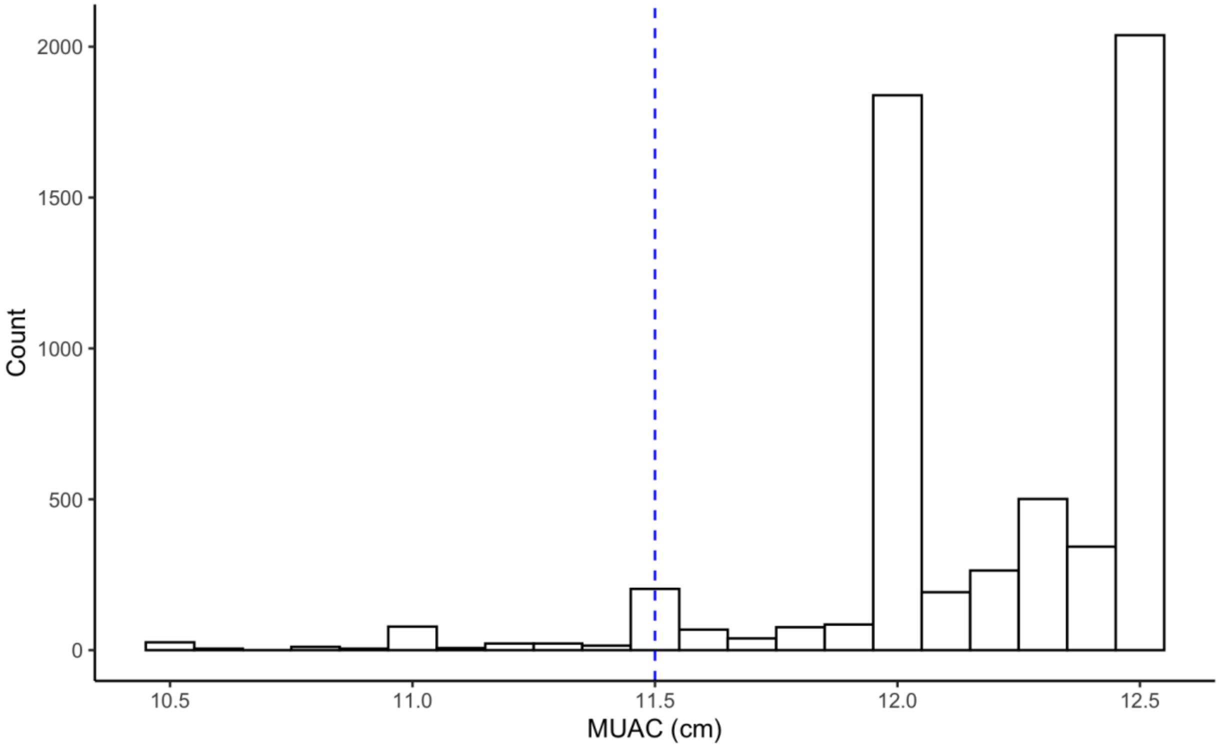

**Supplementary Table 1. Model outcomes by different bandwidth ranges**

| Range | Mean MUAC at 6 months after screening (95% CI) | P-value | Mortality Odds Ratio (95% CI) | P-value |
|---|---|---|---|---|
| **10.5-12.5 cm** | 0.08 (-0.24 – 0.39) | 0.62 | 0.88 (0.09 – 8.83) | 0.91 |
| **9.5-13.5 cm** | -0.14 (-0.35 – 0.08) | 0.22 | 1.12 (0.24 – 5.22) | 0.89 |
| **8.5-14.5 cm** | -0.22 (-0.42 – -0.02) | 0.03 | 42.54 (1.71 – 1057) | 0.02 |

**Supplementary Table 2. Model outcomes after adjusting baseline age, distance to CSPS, number of children (under 5) in household and treatment arm**

| Range | Mean MUAC at 6 months after screening (95% CI) | P-value | Mortality Odds Ratio (95% CI) | P-value |
|---|---|---|---|---|
| **IK-selected bandwidth** | 0.21 (-0.23 – 0.64) | 0.35 | 1.01 (0.03 – 34.15) | 0.99 |
| **10.5-12.5 cm** | 0.08 (-0.24 – 0.39) | 0.64 | 0.82 (0.08 – 8.40) | 0.87 |
| **9.5-13.5 cm** | -0.11 (-0.33 – 0.11) | 0.31 | 1.10 (0.24 – 5.12) | 0.90 |
| **8.5-14.5 cm** | -0.20 (-0.40 – 0.01) | 0.06 | 0.02 (0.00 – 1.13) | 0.06 |

**Supplementary Table 3. Model outcomes adding a squared term to the regression model**

| Range | Mean MUAC at 6 months after screening (95% CI) | P-value | Mortality Odds Ratio (95% CI) | P-value |
|---|---|---|---|---|
| **IK-selected bandwidth** | -0.07 (-0.83 – 0.69) | 0.85 | 0.00 (0.00 – 2,485,404) | 0.20 |
| **10.5-12.5 cm** | 0.13 (-0.43 – 0.69) | 0.66 | 0.89 (0.01 – 79.10) | 0.96 |
| **9.5-13.5 cm** | 0.03 (-0.34 – 0.40) | 0.88 | 0.60 (0.04 – 8.93) | 0.71 |
| **8.5-14.5 cm** | 0.13 (-0.20 – 0.46) | 0.43 | 6.12 (0.42 – 88.90) | 0.19 |
